# Impact of adherence and stringency on the effectiveness of lockdown measures: a modelling study

**DOI:** 10.1101/2024.05.23.24307781

**Authors:** Joren Brunekreef, Alexandra Teslya, Vincent Buskens, Hendrik Nunner, Mirjam Kretzschmar

**Author notes:** These authors contributed equally to this work (Joren Brunekreef, Alexandra Teslya).

## Abstract

During the COVID-19 pandemic, lockdowns were a widely used strategy to reduce disease transmission. However, there was much debate about the optimal level of strictness and duration of lockdowns. This study considers how lockdowns impact public health opinions, which in turn influence adherence to and effectiveness of these measures. We developed an agent-based simulation model to make a theoretical exploration of the impact of health-related opinions on the effectiveness of lockdowns in controlling disease spread. We simulated these dynamics within a hypothetical population connected via a simplified contact network (Watts-Strogatz), incorporating feedback loops between disease prevalence, opinions, and behaviour, including ‘lockdown fatigue’. We explored different scenarios of lockdown implementation in our hypothetical population network by varying a threshold value of prevalence when a lockdown is initiated and the stringency of the lockdown. Our qualitative findings imply that quickly imposing a lockdown with high stringency is the most effective at reducing infection spread, provided that there is a certain degree of adherence to the lockdown among the population. Furthermore, stricter lockdowns minimize fatigue with respect to the imposed measures, since the duration of a lockdown is shorter on average in this scenario. These theoretical results imply that such lockdown policies might therefore be a beneficial, high-impact tool in containing epidemic spread, especially when supplemented by information interventions maintaining the adherence to lockdown measures.

**Author summary:** Lockdowns were a widely used strategy to curb disease transmission during the COVID-19 pandemic. However, the effectiveness of such measures depends on the population’s adherence to the regulations. Long and strict lockdowns may lower this adherence as the population experiences ‘fatigue’ with the regulations. In our study, we model the interplay between lockdowns, disease transmission, and health-related opinions in a population network.

We tested various lockdown scenarios by altering strictness of measures on the one hand, and lockdown initiation times based on disease prevalence rates on the other hand. Our findings show that swift, stringent lockdowns are the most effective in reducing infections, particularly with strong public compliance. Additionally, stricter lockdowns tend to reduce fatigue as they are generally shorter.

These results highlight that prompt, rigorous lockdown policies, supported by strategies to maintain public adherence, are most efficient in controlling epidemics. Furthermore, effective communication to ensure community cooperation may enhance the success of these measures.

## Introduction

During the SARS-CoV2 pandemic, which emerged at the end of 2019 and quickly spread worldwide, combinations of non-pharmaceutical interventions (NPIs), including government-imposed lockdowns, were used to mitigate the spread of the virus. Before vaccines became available at the end of 2020, NPIs were the only public health intervention available to curb the pandemic. Since then, many studies have been published in which the effectiveness of NPIs has been investigated (e.g. [1–4]). However, it is difficult to extract information about the effectiveness of specific measures, as they were usually implemented as a part of a larger set of measures. Lockdowns were the most stringent type of NPIs used. Aiming to reduce the contact rates as much as possible, the mobility of the population was significantly restricted and large gatherings were not allowed [5]. Lockdowns were imposed by governments and usually had precise start and end dates, chosen according to the pandemic situation at the time. The exact realization of lockdowns in different countries varied in terms of chosen measures and their implementation. Thus, to facilitate comparison, an index quantifying the stringency of measures on the population level was developed [6]. This stringency index is computed as the mean score of nine different ordinal metrics that indicate the extent to which a certain lockdown policy has been imposed. Examples of such policies are school closures, cancellation of public events, and stay-at-home requirements.

Policymakers must consider several critical aspects when developing a strategic plan for implementing a lockdown. These considerations include determining the optimal timing for initiating the lockdown based on the prevalence or incidence of new cases, deciding the level of stringency for the lockdown measures, and establishing the appropriate duration. These factors not only affect the epidemiological outcomes of the lockdown but also have significant implications for the economic burden, mental health impacts, and potential social unrest within the population. While early implementation and prolonged duration of lockdowns can significantly reduce case numbers, these benefits might be offset by negative consequences, such as diminished adherence to the lockdown measures [**?**, 7–10]. While many studies have attempted to quantify the epidemiological effects of NPIs and lockdowns, few have considered the impact of adherence of the population on the effectiveness and duration of lockdowns [11].

In this study, we use an agent-based model to explore how adherence to lockdown measures affects epidemiological outcomes, such as the number of new cases and the population’s attitude towards protective measures. In our model, the decision to initiate or end a lockdown is based on specific epidemic thresholds. The population in the model is connected through a network of social contacts, allowing for the spread of both health-related opinions and the infectious disease. Essentially, our model represents a complex system where opinion dynamics and infection spread coexist in a connected network. Our approach builds on earlier work [2, 12, 13], where we investigated the interplay between opinion and epidemic dynamics. In the present work, we studied the effect of different aspects of lockdown implementation on epidemic outcomes, population health opinion dynamics, as well as duration and number of lockdowns taking place within the first year of an outbreak.

## Materials and methods

### General description of the model

We consider the emergence of an airborne infectious disease in a closed population. Transmission of the disease requires close physical contact for an extended amount of time (such as standing within 1.5 meters of each other for at least 15 minutes). In the context of the disease, members of the population hold one of two mutually exclusive health-related opinions: health-positive and health-neutral. These opinions affect the behavior, which in turn changes the individual probability of contracting the infection. Self-protective behaviour includes both self-applied measures (such as hand washing) as well as the contact pattern of individuals (i.e., limiting contacts to avoid infection). The basic assumptions concerning the interaction between epidemic and health opinions dynamics in our agent-based model draw from the framework outlined in [12]. In this work, the authors developed a deterministic model incorporating feedback between epidemic dynamics and competing health opinions and used it to study the impact of contact rates and assortativity by opinions on epidemic dynamics. We extended the model by incorporating government-imposed physical distancing measures and their effects on health opinion dynamics.

In the model, individual health-related opinions change over time, driven by several factors. Switching of opinions occurs as a result of exchanging information with peers, such that the more common an opinion is in the population, the higher the switch rate to this opinion. The presence of the disease is not required for both opinions to persist. During an outbreak, the adoption of a health-positive opinion leading to self-protective behavior is positively correlated with the prevalence of the disease, thus creating a feedback loop between the health opinion dynamics and disease dynamics. When an outbreak ends, the switch rate to the health-positive opinion is reduced to the pre-pandemic level. We assume that opinion dynamics and epidemic dynamics occur on roughly similar time scales, such that steady state approximations cannot be used. Finally, we consider the possibility of a temporary large-scale societal lockdown when the disease prevalence rises above a certain predefined threshold. For the duration of a lockdown, the average rates of close physical interactions experienced per individuals are reduced for the entire population. In addition to impacting the transmission of the disease, a lockdown influences health-related opinion dynamics by inducing *lockdown fatigue* [7–9, 14], which may increase the adoption of the health-neutral opinion and subsequently the overall decrease in adoption of self-protective measures [10]. Lockdowns are lifted when the disease prevalence drops below a second predefined threshold. For simplicity, following the lifting of the lockdown, the switch rate to the health-neutral opinion is immediately re-set to the pre-lockdown level.

### Model description

We summarize the key features of the model here, and refer the reader to the S1 p for the technical formulations. The model that we use is a stochastic agent-based network model, where nodes of the network correspond to individuals and edges indicate that the individuals linked by an edge engage in regular contact with one another. These contacts can be of a *physical* nature, meaning any type of (social) interaction within close proximity for a minimal extended duration of time, or of an *informational* nature, where only information is exchanged by e.g. text messages, email, or phone calls. We call two individuals *peers* if they are connected by an edge. Disease transmission can occur exclusively through physical interactions, whereas opinion switches are driven by the informational interactions.

At the start of the simulation, we generate a small-world population network by use of the Watts-Strogatz algorithm [15], and all edges of the resulting network are considered to be both of a physical and informational nature. We chose the Watts-Strogatz model as it generates networks capturing high clustering and short average path lengths, commonly referred to as ‘small-world’ properties. These characteristics are often observed in social networks [16] and are relevant for modelling both localized transmission and broader information spread. We acknowledge that this choice amounts to a strong simplification in which several features of real-world networks are not captured. However, the current work can be seen as a computationally tractable starting point, and the effect of using more realistic network structures could be explored in future work. For more details about the properties of the networks in the simulations, we refer to S1 p.

The network is static in all scenarios, but the nature of the contact edges can change when a lockdown is initiated: for the duration of a lockdown, the *physical* interaction along certain edges in the network is temporarily disabled, preventing the transmission of disease through these contacts. We elaborate on the exact implementation of this lockdown mechanism later in this section.

### Disease dynamics

The model population has fixed size *N*, and we omit considerations of birth and death due to the relatively short time horizon of our study, approximately one year. The disease dynamics are modeled using an SIR framework, where individuals in the population are categorized into three states: *susceptible* (S), *infected* (I), or *recovered* (R). A susceptible individual can get infected through a physical contact (symbolized by a network edge) with an infected individual. The likelihood for an individual to get infected at any moment in time depends on

1. the physical contact rate *c*_phys_ of the edge,
2. the number *n_I_* of the individual’s infected peers, and
3. the infection probability per contact *ɛ*.

We set the physical contact rate *c*_phys_ of all edges to the same value for simplicity. Once individuals are infected, they naturally recover, on average, after 1*/γ* weeks (with an exponential distribution of duration of infectiousness). Following the recovery, individuals are immune to re-infection. At the start of an outbreak, all individuals in the population are susceptible.

Each individual in the population holds one of two mutually exclusive health-related opinions. The individuals with *health-positive* opinion (denoted⊕) modify their behavior with the effect of reducing the probability of contracting an infectious disease, whereas the individuals of *health-neutral* opinion (denoted⊗) do not modify their behavior and hence are more susceptible to infection. Therefore, the infection probability *ɛ* can depend on opinion status, and we denote these probabilities by *ɛ*^⊕^ and *ɛ*^⊗^. We make the simplifying assumption that these behavioral differences affect only susceptibility and do not influence the infectivity of the infected individual.

### Lockdowns

The central element of our simulation setup is the lockdown mechanism. A lockdown is immediately implemented when the disease prevalence exceeds a threshold initiation value *f_s_*. As briefly mentioned before, lockdowns are effectuated by cutting the physical interaction along certain edges in the network (the informational interaction of those edges remains in place). This cutting of physical edges occurs in a probabilistic manner, where the probability for any edge to be cut is chosen as follows.

When at least one of the individuals connected by an edge is of a health-positive opinion, the cut probability is equal to *q*, where 0 *≤ q ≤* 1. This model parameter *q* is called the lockdown *stringency*, with *q* = 1 corresponding maximal stringency and *q* = 0 being equivalent to a no-lockdown scenario. When an edge connects two health-neutral individuals, the cut probability is *q · α*, where 0 *≤ α ≤* 1 is the lockdown *adherence* parameter. Therefore, when adherence is maximal (*α* = 1) the cut probability is equal for any edge in the network. However, when the adherence is below 1, physical interaction edges between two health-neutral individuals are less likely to be cut than edges involving at least one health-positive individual.

This models the assumption that health-neutral individuals are more likely to be non-adherent to health-related measures, including lockdown regulations. Since meeting up in person during a lockdown requires non-adherence from both parties involved, we opted to invoke the reduced cut probability *q · α* only when both individuals connected by an edge are of health-neutral opinion.

### Opinion dynamics

We distinguish three distinct processes that contribute to opinion switching in the model.

Firstly, switching of opinion occurs through communication with peers holding the opposite opinion, and the likelihood for this switch to occur depends on

1. the informational contact rate *c*_inf_ between individuals,
2. the number of peers *n*_op_ of opposing opinion, and
3. the opinion switch probability per contact *p*.

Similar to our approach for physical contacts, we take the informational contact rate *c*_inf_ to be identical for all edges.

Secondly, switching from a health-neutral to a health-positive opinion (a ⊗ → ⊕ switch) is amplified by an increase in disease prevalence in the population. This is implemented in the model via a dependence of the switch rate on the global prevalence *P*. Although we cannot expect all agents to have perfect knowledge of the precise prevalence at any given point in time, the true prevalence is a good first approximation of the ‘believed’ prevalence.

Thirdly, and finally, lockdown-fatigue can amplify an opinion switch ⊕ → ⊗ with the likelihood of this switch depending on

1. the time *τ* since the start of the current lockdown and
2. lockdown stringency as captured by parameter *q*.

Note that we do not include other ‘global’ opinion switching mechanisms, such as effects of mass media dissemination.

We summarize the model parameters in Table 1, and refer to S2 p for a description of choice of model parameters and model calibration. Here we selected the values for epidemiological parameters to model an acute immunity-inducing disease similar to COVID-19 or influenza. To bring the interplay between the epidemic dynamics, opinion dynamics and public health intervention measures to the forefront, we did not consider a possibility of waning immunity or disease-induced mortality.

**Table 1.**
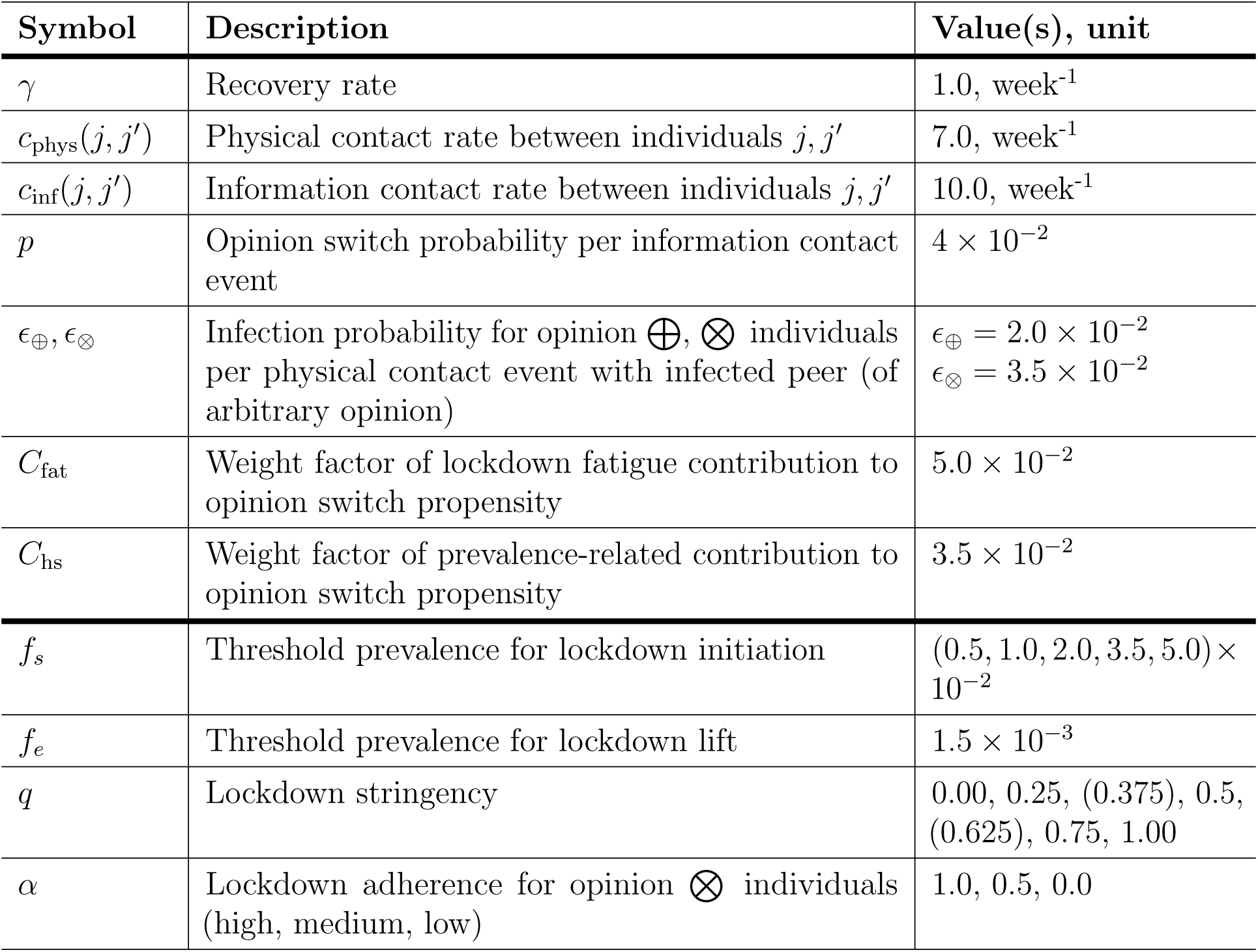
Model parameters.

| Symbol | Description | Value(s), unit |
| --- | --- | --- |
| $\gamma$ | Recovery rate | 1.0, week<sup>-1</sup> |
| $c_{\text{phys}}(j, j')$ | Physical contact rate between individuals $j, j'$ | 7.0, week<sup>-1</sup> |
| $c_{\text{inf}}(j, j')$ | Information contact rate between individuals $j, j'$ | 10.0, week<sup>-1</sup> |
| $p$ | Opinion switch probability per information contact event | $4 \times 10^{-2}$ |
| $\epsilon_{\oplus}, \epsilon_{\otimes}$ | Infection probability for opinion $\oplus, \otimes$ individuals per physical contact event with infected peer (of arbitrary opinion) | $\epsilon_{\oplus} = 2.0 \times 10^{-2}$<br>$\epsilon_{\otimes} = 3.5 \times 10^{-2}$ |
| $C_{\text{fat}}$ | Weight factor of lockdown fatigue contribution to opinion switch propensity | $5.0 \times 10^{-2}$ |
| $C_{\text{hs}}$ | Weight factor of prevalence-related contribution to opinion switch propensity | $3.5 \times 10^{-2}$ |
| $f_s$ | Threshold prevalence for lockdown initiation | $(0.5, 1.0, 2.0, 3.5, 5.0) \times 10^{-2}$ |
| $f_e$ | Threshold prevalence for lockdown lift | $1.5 \times 10^{-3}$ |
| $q$ | Lockdown stringency | 0.00, 0.25, (0.375), 0.5, (0.625), 0.75, 1.00 |
| $\alpha$ | Lockdown adherence for opinion $\otimes$ individuals (high, medium, low) | 1.0, 0.5, 0.0 |

We initialized the population with equal proportions of health-positive and health-neutral opinions, randomly distributed over the network. This was followed by an “opinion burn-in” phase, where opinions circulate in the network in the absence of disease in order to settle their distribution to the quasi-steady state emerging from the definition of the model. To exhaustively explore the impact of the feedback between epidemiological variables, opinion dynamics and NPIs, we have considered various scenarios which were captured by varying key parameters across plausible intervals. The range for the stringency parameter was chosen so that restrictions varied from minimal to complete, effectively eliminating all contacts. The thresholds for initiating lockdown based on infection prevalence varied from 0.5% to 5%. Finally, we considered three levels of adherence among individuals with health-neutral opinions: high (1), medium (0.5), and low (0). For each combination of parameter values, we performed 500 distinct simulation runs with a runtime of 50 weeks. We chose this time frame so the assumption of the permanence of the state of the population, such as turn around and availability or lack of biomedical interventions, remains plausible.

Preliminary simulations showed a reversal of epidemiological outcomes around the stringency *q* = 0.25 point for the high-adherence (*α* = 1.0) scenario. In order to explore this regime in more detail, we added two nearby values of *q* for this scenario to our list of parameter combinations for full simulation runs.

Each simulation run started in a state without a lockdown in a fully susceptible population, with 3 index infection cases at randomly selected nodes in the network. As a consequence of this small number of index cases, in a small number of simulation runs, the disease stochastically goes extinct. These runs were also included in the aggregated results. We compared scenarios with lockdowns to a baseline scenario, where a lockdown is not imposed, corresponding to the stringency parameter set to *q* = 0. We summarized the runs by calculating mean values of the following outcomes at the end of each simulation, i.e., at 50-week mark:

- Relative final outbreak size (RFOS). The ratio of the final outbreak size and the mean final outbreak size for the lockdown-free scenario (*q* = 0). If the relative final outbreak size is smaller than 1, lockdown measures have led to an overall reduction in disease spread, whereas a relative final outbreak size larger than 1 indicates that, due to the lack of adherence to lockdown measures, the final number of infections was higher than in the baseline scenario.
- Final proportion of health-neutral (⊗) individuals (FPHN).
- Cumulative duration and frequency of lockdowns.

The final outbreak size is defined as the proportion of the population that has contracted the disease — that is, the proportion of the total population who are either infected or recovered when the simulation ends.

Finally, to illustrate a range of features of the time course of a pandemic in different scenarios, we present the time series of several individual simulation runs.

The simulation code was written in MATLAB (R2023a) [17], and is publicly available on GitHub [18]. Wolfram Mathematica 13.0 [19] was used for data analysis and generating figures.

## Results

### Disease and opinion outcomes

#### Low lockdown stringency

For low lockdown stringency (e.g., *q* = 0.25), we find that the final outbreak size is comparable to or worse than that of a lockdown-free scenario (*q* = 0), regardless of the lockdown initiation threshold prevalence *f_s_* or the adherence to lockdown regulations by health-neutral individuals *α* (Fig. 1**a-c**). Additionally, most individuals have adopted a health-neutral (⊗) opinion by the end of the year (Fig. 1**d-f**), making the population more susceptible and therefore more vulnerable overall to future epidemic outbreaks.

**Fig 1.**
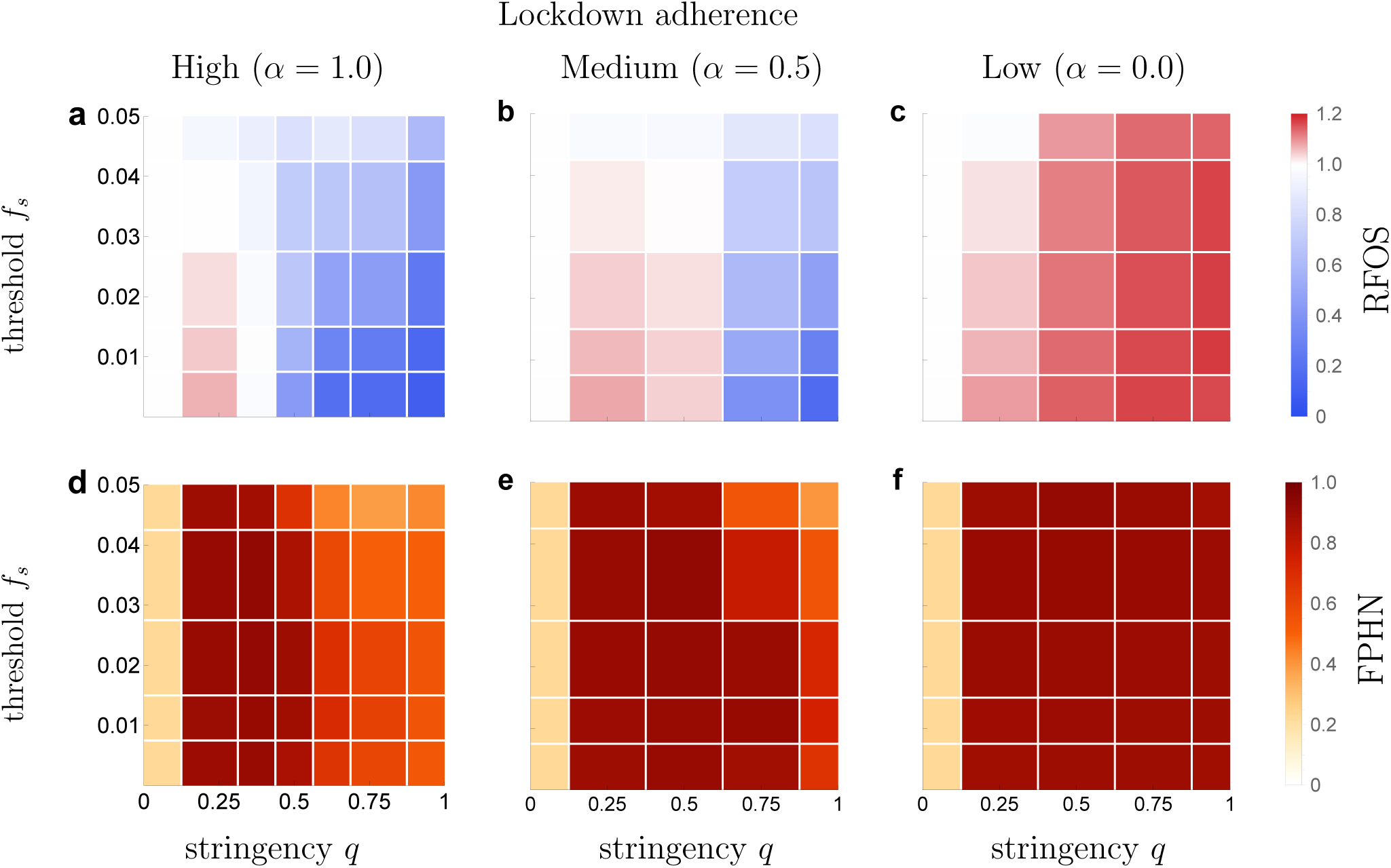
Effects of the lockdown on the outbreak size and distribution of opinions one year after outbreak starts. Mean final outbreak size relative to mean outbreak size for scenarios without lockdowns (RFOS, **a-c**) and mean final proportion of health-neutral individuals (FPHN, **e-f**) at 50-week time mark. All outcomes are shown in relation to lockdown stringency *q* and lockdown initiation threshold *f_s_* for varying degrees of lockdown adherence *α*.

#### Medium lockdown stringency

For lockdowns with medium stringency (e.g., *q* = 0.5), the outcome depends on the threshold prevalence *f_s_* and the adherence to lockdown measures *α*. If the adherence to lockdown measures is high (e.g., *α* = 1.0), we observe that the final outbreak size can be significantly reduced by imposing lockdown measures (Fig. 1**a**). This effect increases when the lockdown initiation threshold prevalence is reduced — corresponding to lockdowns initiated early in the outbreak. On the other hand, this policy causes a large proportion of the population to adopt a health-neutral opinion (Fig. 1**d**).

When the adherence to lockdown measures is low or medium (*α* = 0.0, 0.5), lockdown measures have a net detrimental effect on both the final outbreak size and the proportion of health-neutral individuals at the 50-week mark (Fig. 1, **b**, **c**, **e**, **f**). Furthermore, this effect grows stronger as the prevalence threshold *f_s_* is reduced.

#### High lockdown stringency

The dynamics for scenarios with strict lockdowns (*q >* 0.5) exhibit a strong dependence on both the initiation prevalence threshold *f_s_* and the adherence *α*. When adherence is medium or high (*α* = 0.5, 1.0), we find that lockdown measures can be very effective at reducing the final size of infection numbers over the initial 50 weeks. Furthermore, the results show that in this scenario it is possible to ensure that a large proportion of the population holds a health-positive (⊕) opinion at the 50-week mark: on the one hand by imposing strict lockdowns, and on the other hand by initiating lockdowns at a *higher* critical prevalence. However, this latter strategy comes at the cost of a comparatively higher final outbreak size.

In the scenario where adherence is low (e.g., *α* = 0.0), lockdowns are detrimental with respect to every metric shown in Fig. 1, regardless of the lockdown initiation threshold *f_s_*. At the mark of 50-weeks, the population is in a state where almost every individual holds a health-neutral opinion, and the mean final outbreak size can exceed the lockdown-free level by as much as 20%.

### Duration and frequency of lockdowns

In addition to outcomes pertaining to epidemic and opinion dynamics, we investigated emerging duration and frequency of lockdown measures that are imposed on the population in each scenario. We present the results in Fig. 2.

**Fig 2.**
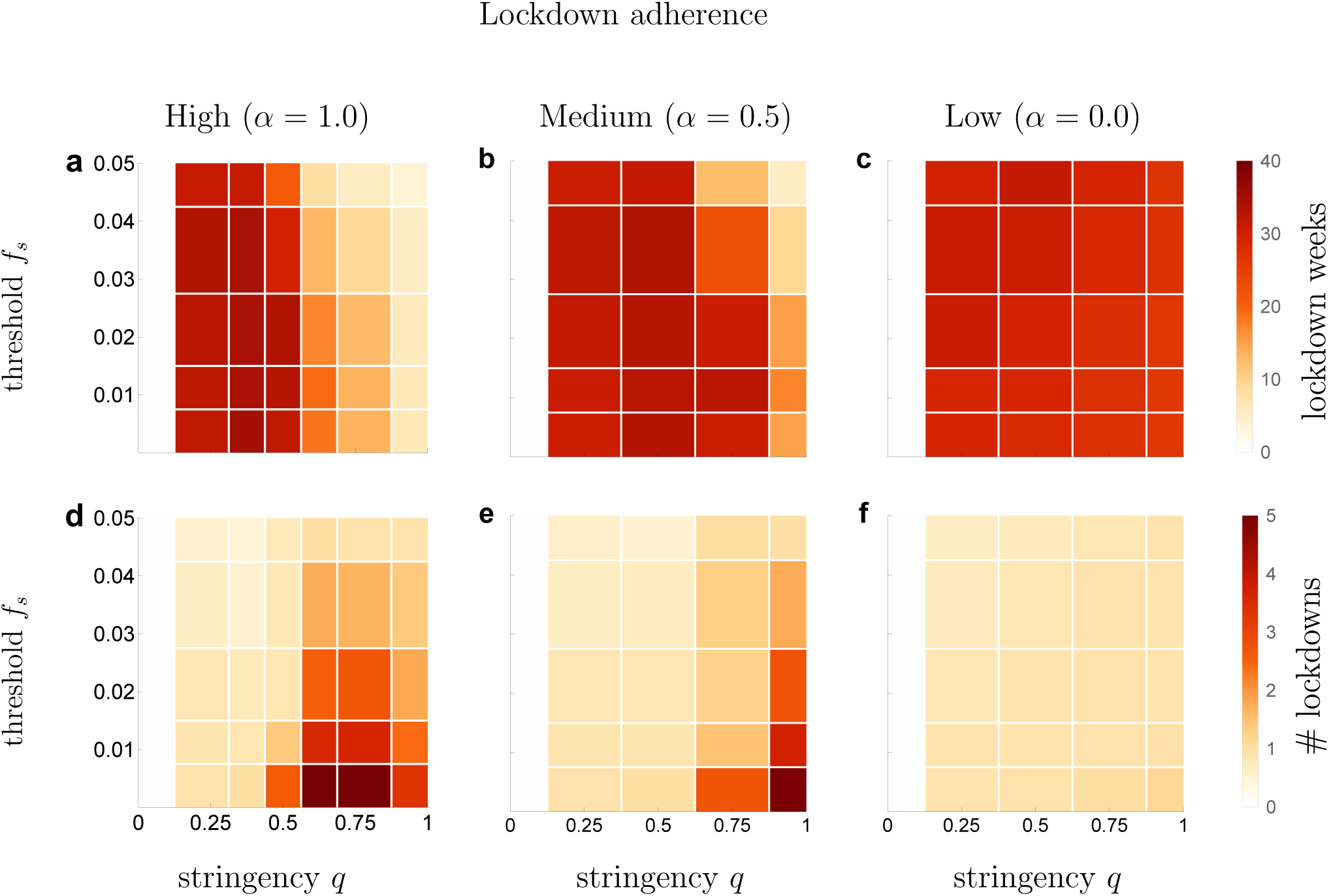
Dependence of the overall lockdown duration and the number of lockdowns on the stringency of lockdowns, initiation threshold and adherence. **a-c** Mean number of weeks spent in lockdown; **d-f** mean number of distinct lockdowns. White region on the left side in each plot corresponds to stringency *q* = 0, which describes a scenario where lockdowns are not imposed.

#### Low and medium lockdown stringency

With a low or medium lockdown stringency of 0.25 *≤ q ≤* 0.5, regardless of lockdown adherence, we find that on average the measures fail to reduce the prevalence below the lockdown-lifting threshold *f_e_* within the simulated 50-week period. As a result, the population will spend the majority of this period in a single, long lockdown, starting from the moment when the disease prevalence initially increases to the lockdown-initiation threshold *f_s_*. Interestingly, the population tends to spend more time in lockdown when lockdown adherence is high (*α* = 1.0) than when it is low (*α* = 0.0).

#### High lockdown stringency

When lockdown measures are strict (*q >* 0.5), in a setting with medium to high adherence to the lockdown (*α* = 0.5, 1, Fig. 2**a**, 2**b**, 2**d**, 2**e**), the outcomes exhibit a strong dependence on the lockdown initiation threshold *f_s_*. More specifically, as the initiation threshold decreases the duration and the number of lockdown increases. One possible explanation for this is that for a low threshold, such as *f_s_* = 0.005, a strict lockdown drives infection rates down quickly, causing the prevalence to drop below the lockdown-lifting threshold *f_e_*. As a result, the lockdown ends while the majority of the population is still susceptible, and the duration of the lockdown is too brief to promote a shift towards a health-neutral opinion. Subsequently, if the disease does not go extinct, infection numbers may start rising again, soon leading to a new lockdown. As a result, the mean number of lockdowns and their total duration over the full 50-week period are high. If the lockdown adherence is low (e.g., *α* = 0.0, Fig. 2**c** and 2**f**), the level of lockdown stringency and adherence of individuals to the lockdown do not have an effect on the duration of the lockdown or the number of times it is initiated. In this scenario, the population will experience a single lockdown with a long duration of approximately 30 weeks or more. This lockdown does decrease infection transmission, but due to the lack of adherence by a part of the population, the transmission is still ongoing, keeping prevalence above the lockdown termination threshold.

### Duration of lockdown versus final outbreak size

To show the relationship between the total time spent in lockdown (*T*_ld_) and the relative final outbreak size across various scenarios, we plotted these quantities against each other for all parameter combinations and all simulation runs in Fig. 3. This plot also shows the stochastic variability in outcomes per parameter combination, color-grouped by the lockdown stringency parameter *q*. The three distinct graphs correspond to three settings of adherence to lockdown, high, medium, and low with *α* = 1.0, 0.5, 0.0, respectively. The points in the lockdown-free scenario (*q* = 0) cluster around a relative final outbreak size of 1, and all have *T*_ld_ = 0. When the relative final outbreak size exceeds 1, lockdowns have led to a higher final outbreak size than could have been expected in the absence of lockdown measures. In this analysis, we have fixed the lockdown initiation prevalence at *f_s_* = 0.005 in all simulation runs. For analogous figures showing the final proportion of health-neutral individuals, we refer to S3 p.

**Fig 3.**
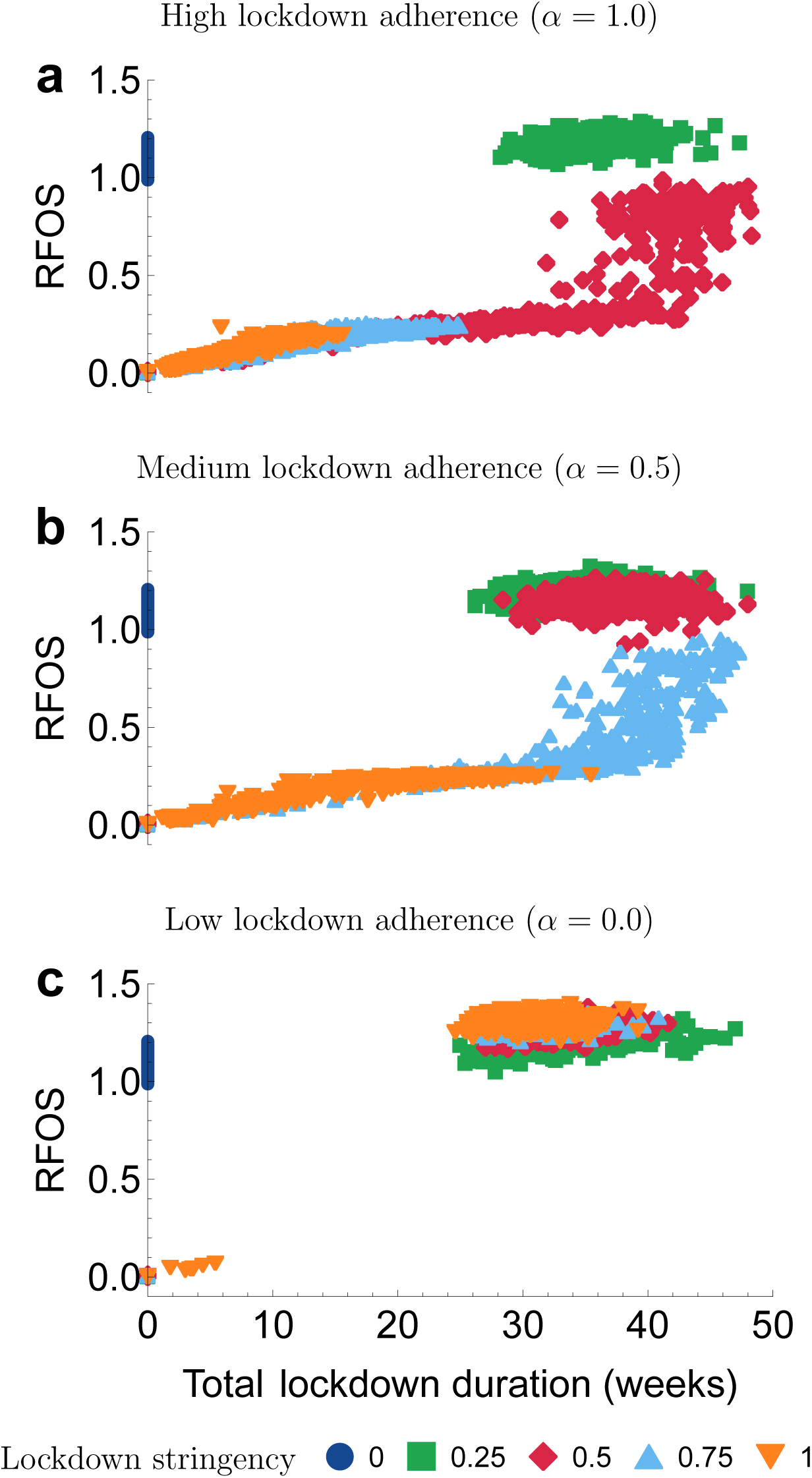
Spread of outbreak sizes and lockdown duration. The panels show relative final outbreak size (RFOS) versus the total lockdown duration, across various levels of lockdown adherence parameter *α* = 1.0, 0.5, 0.0. The outbreak size is reported relative to the mean final outbreak size in the scenario where no lockdown is implemented (*q* = 0). Each marker corresponds to a single simulation run. The threshold prevalence for lockdown initiation is fixed at *f_s_* = 0.005.

The outcomes in the low-stringency lockdown scenario (*q* = 0.25, green squares) always cluster in the top right of the graph, regardless of the adherence *α*. The outcomes of this scenario exhibit long lockdowns with high final outbreak sizes.

For a medium lockdown stringency (*q* = 0.5, red diamonds), depending on the level of adherence *α*, spread of both the relative final outbreak size and the total time spent in lockdown can range over a wider region. When adherence is high (*α* = 1.0, Fig. 3**a**), the relative final size is dispersed over a large interval, varying from nearly 1, indicating minimal reduction, to substantial reductions in the final size. Notably, even when the final size is small, the duration spent in the lockdown can still be significant (20 or more weeks). When the relative final outbreak size is close to 1, the total duration of lockdown spans approximately 40 weeks or more. A small number of points are found in the bottom left, where the total lockdown duration and relative final outbreak size are low. This corresponds to stochastic extinction following the first lockdown. Most outcomes, however, are characterized by longer total lockdown duration *T*_ld_. When adherence is lower (*α* = 0.0, 0.5, Fig. 3**b** and 3**c**) all points for medium-stringency lockdown scenarios are located at the top right corner, signifying that while a large amount of time was spent in lockdown, the epidemiological outcomes are equal or worse compared to a lockdown-free scenario.

This pattern continues as the stringency of the lockdown, *q*, increases. The higher the value of *q*, the more points remain in the region where the duration of lockdown is short and relative final outbreak size is low, but a lower adherence (*α* = 0) almost always has the effect of leading to long lockdown duration and higher relative final outbreak size.

### Representative simulation trajectories

The results we have shown so far captured the state of the population at the end of the simulation run, i.e. the state of the population after *t* = 50 weeks. Studying the time evolution of individual simulation runs provides insight into the mechanisms that lead to the final outcomes we presented above. In Fig. 4, we show time series of the prevalence (left column) and proportion of individuals with a health-neutral (⊗) opinion (right column), for several combinations of the simulation parameters (*q, α, f_s_*).

**Fig 4.**
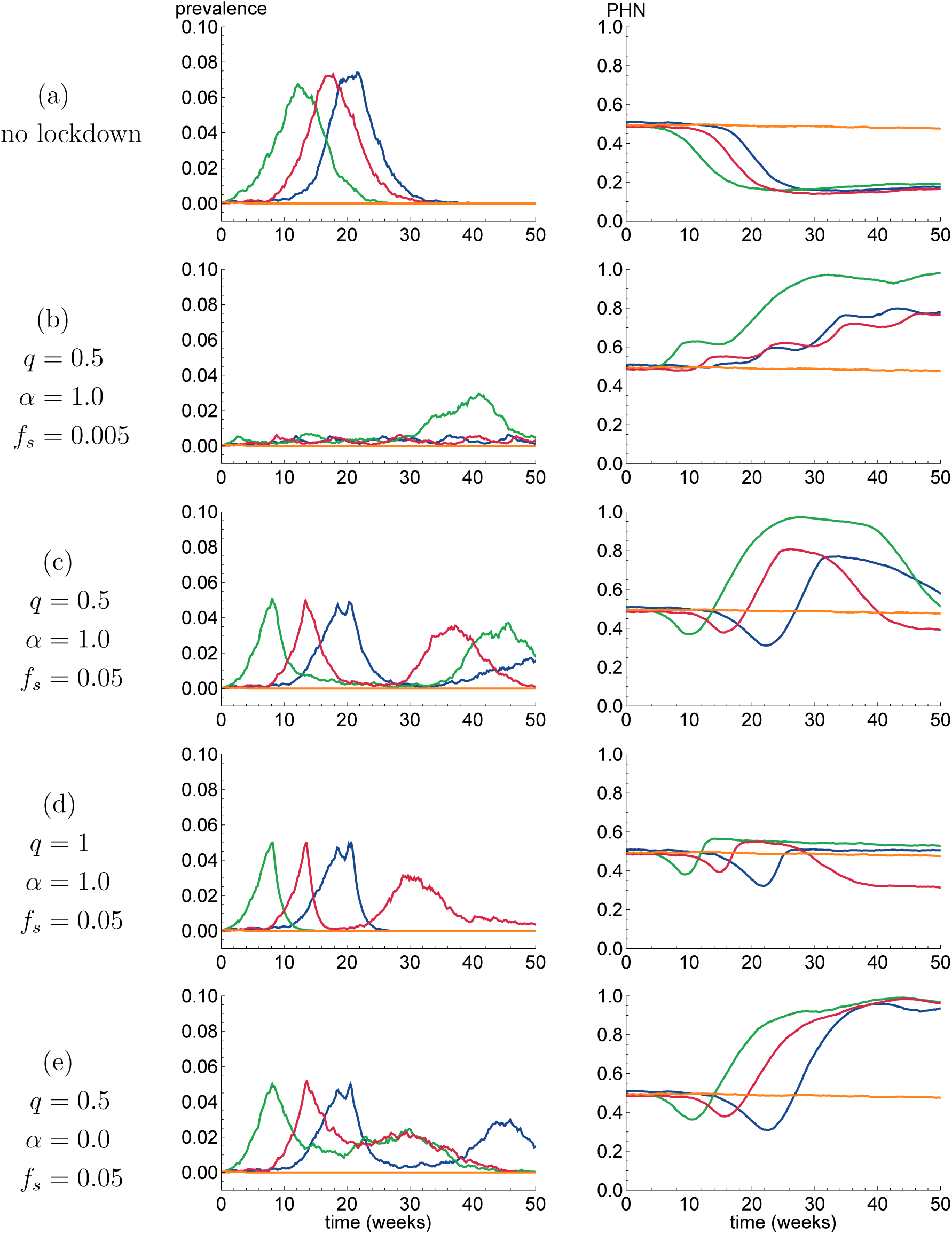
Four individual simulation runs (distinguished by four colors) are shown for five sets of parameter values (each set in one row). The left column shows prevalence over time, and the right column shows the proportion of health-neutral opinion (PHN) in the population. The nearly constant orange trajectory is an example of a situation where the infectious disease goes extinct shortly after it is introduced into the population.

For each parameter set, we plot the results of four distinct simulation runs. Note that the initial behavior of the trajectories is *independent* of the choice of (*q, α, f_s_*), since the population always starts in a state without lockdowns. These three parameters start to affect infection transmission dynamics once the prevalence has risen above the threshold *f_s_* for the first time, and a lockdown is initiated.

In order to simplify the comparison between the different parameter choices, we used colors to indicate simulation runs that have *identical* initial behavior. That is, each color corresponds to a specific choice of seed for the pseudo-random number generator used by the simulation. As pointed out above, all simulations are equivalent before the first lockdown has started, so equal seeds will produce identical initial trajectories in the rows of Fig. 4.

#### Lockdown-free scenario

The baseline scenario is the situation where a lockdown will not be imposed (refer to Fig. 4**a**), which is equivalent to a scenario with lockdown with stringency *q* = 0. We observe typical SIR epidemic dynamics: prevalence rises quickly after the infectious disease is introduced at *t* = 0. As more people recover from the disease and the number of susceptible individuals decreases, infection rates decrease and the prevalence eventually drops to zero. Furthermore, the high prevalence induces increased switching to a health-positive opinion causing reduction in the proportion of individuals holding a health-neutral opinion for an extended period of time.

In one of the trajectories (in orange), the epidemic does not take place since the infection goes extinct within a few weeks. In this scenario, prevalence levels do not become high enough to promote increased switching from health-neutral to health-positive opinion, and as a result the distribution of health-related opinions remains close to its initial value of 0.5.

#### Medium stringency, high adherence, low initiation threshold

In the scenario where *q* = 0.5, *α* = 1.0, and *f_s_* = 0.005 (row (b) in Fig. 4), lockdowns are initiated at very low prevalence levels; they have medium stringency and the adherence to them is strong. Since in such conditions the disease does not spread easily, lockdowns are effective at keeping prevalence at low levels. As a result, the prevalence soon decreases below the lockdown-lifting threshold *f_e_* = 0.0015 and the lockdown ends. However, the disease is still present in the population and most of the population is susceptible to infection. Subsequently, the number of infected individuals can start rising again, leading to a new lockdown. Each time a new lockdown is initiated, the number of health-neutral individuals is seen to increase. Even though adherence to the lockdown is high, health-neutral individuals do not engage in self-protective measures and are therefore more likely to get infected. In the green trajectory, it is clear that this dynamic can have a detrimental effect on epidemic control in the long run: after week 30, a substantial part of the population is of a health-neutral opinion and the disease can spread even during a lockdown.

#### Medium stringency, high adherence, high lockdown initiation threshold

In the scenario where *q* = 0.5, *α* = 1.0, and *f_s_* = 0.05 (row (c) in Fig. 4), the lockdown is initiated at a higher prevalence and therefore, comes in effect later. Once the lockdown is initiated, some time later prevalence decreases until it drops to a point where the lockdown measures are lifted again. The cycle then repeats, although the second prevalence peak is lower than the first. The opinion distribution graphs in the right column show that initially there is a tendency to switch from health-neutral to health-positive opinion as disease prevalence increases, but this is subsequently counteracted by lockdown fatigue and, as a result, individuals start to switch to the health-neutral opinion. In this scenario, individual lockdown periods can last longer compared to those expected with a lower initiation threshold *f_s_* (refer to Fig. 2**a**), potentially causing a large fraction of the population to adopt a health-neutral opinion as a result of longer lockdowns. However, with more individuals holding the health-neutral opinion, the overall population becomes more susceptible, making a large second wave of infections possible. This secondary wave may in turn prompt a reversal in health opinion dynamics with a large fraction of the population adopting a health-positive opinion.

#### High stringency, high adherence, high threshold lockdown initiation

In the scenario where *q* = 1.0, *α* = 1.0, and *f_s_* = 0.05 (refer to Fig. 4**c**), prevalence dynamics are expected to be qualitatively similar to those shown in row (c), but the lockdowns, due to their increased stringency, are much more effective at quickly stemming infection transmission and therefore, keeping the duration of the lockdowns short. In three of the four plotted trajectories, the lockdowns are sufficiently effective to lead to the disease’s extinction within the time horizon considered, such that no second wave occurs. In this scenario, a small proportion of the population adopts a health-neutral opinion during the lockdown, but its short duration ensures lockdown fatigue remains low and subsequently the opinion distribution returns to the pre-outbreak state. In fact, the only trajectory (in red) where a second wave occurs has a *higher* proportion of health-positive individuals at the 50-week mark. This happens since the second wave does not reach the lockdown initiation threshold prevalence. As a result, the proportion of the population engaging in self-protective measures increases, and the prevalence slowly decreases without the need for further lockdown measures.

#### Medium stringency, low adherence, high threshold lockdown initiation

In the scenario where *q* = 0.5, *α* = 0.0, and *f_s_* = 0.05 (refer to Fig. 4**e**), lockdowns are imposed, but fail to quickly reduce the prevalence to a level low enough for the restrictions to be lifted. Subsequently, long-lasting lockdowns lead to lockdown fatigue causing a large proportion of the population to switch to the health-neutral opinion. This shift results in increased susceptibility of the overall population.

## Discussion

Using an agent-based model, we found that stricter lockdowns could be more effective for epidemic control than less strict ones. This is in line with the findings of a data analysis from 8 European countries which has shown the stringency of lockdowns to be one of the most important factors affecting the spread of SARS-CoV-2 [20]. Similarly, in an analysis of data from Queensland, Australia, Vogler et al. [21] concluded that there was a statistically significant relationship between stringency of measures and case numbers. However, adherence of the population to lockdown measures is essential for the success of the lockdown in terms of preventing new infections and for keeping lockdowns short. Our findings indicate that insufficiently strict lockdowns not only can lead to higher numbers of infections, but also can contribute to increase of lockdown fatigue. Subsequently, this may influence individuals into adopting a health-neutral opinion, wherein they may be less willing to take protective measures. Jointly these dynamics can lead to prolonged cumulative lockdown duration with possibly adverse economic consequences.

The epidemic threshold for initiating a lockdown is another crucial factor. For strict lockdowns, there is a negative association between the prevalence threshold at which lockdown measures come into effect and the duration of the lockdown. Initiating lockdowns at a lower prevalence threshold is projected to extend the cumulative duration of lockdown measures compared to initiating them at a higher threshold, although the former approach prevents more infections. We conclude that the threshold needs to be set where onward transmission is slowed down quickly, thus minimizing the burdens on the health care system and work force, while also ensuring that the time spent under a lockdown is kept short. The optimal threshold varies depending on the specific characteristics of an infection, e.g., generation time distribution, fraction of transmissions, that occur before symptom onset, and morbidity and mortality case ratio. Vogler et al. [21] when discussing the timeliness of measures, conclude that many factors have to be taken into account to determine when a lockdown should best be implemented.

Nevertheless, given a sufficiently strict lockdown, even at high initiation threshold, a lockdown can be expected to decrease the number of new infections compared to the baseline. From a public health perspective, these results suggest that strict lockdown measures can be more effective in controlling the epidemic than incremental, step-by-step measures. Ultimately, strict measures may reduce public health burden (in terms of infections, hospitalizations, and disease-induced mortality) and may have lower economic impact than less strict measures. However, our results suggest that lockdown measures should be accompanied by information campaigns to maintain strong public support and by provision of resources to the population enabling adherence. Communication that promotes health-positive opinions and combats lockdown fatigue is essential to the success of lockdowns as means to control an epidemic.

This study has several limitations. In the model, opinion dynamics are simplified, reducing a continuous realm to two mutually exclusive opinions, with modeling framework based on a previous study [12]. While the network degree distribution and contact rates were derived from literature [22, 23], we did not formally calibrate our model to specific data sets. In terms of infection transmission parameters, we used estimates consistent with SARS-CoV-2 but did not aim to precisely replicate its natural history. The model, despite its simplified approach to infection and opinion dynamics, incorporates many parameters, some of which are not easily derived from the existing literature. We conducted sensitivity analyses on key parameters, while maintaining others constant, rather than exhaustively explore all possible parameter variations.

As we pointed out in the Methods section, the use of only Watts-Strogatz type networks amounts to a strong simplification that was necessary to make this project computationally feasible and to restrict the scope. While these networks capture some features of real-world population networks, such as ‘small-world’ properties, the networks are still too simple to properly resemble the complex structures present in real populations. The influence of using different network topologies on the interplay of opinion and disease dynamics remains an important area for future research.

We also did not include disease-induced mortality or waning immunity in the model. Both are important factors in real-world epidemic response. Mortality, in particular, influences public perception and policy decisions, but in our framework, it closely follows prevalence, which already drives opinion shifts and intervention triggers. Including mortality would therefore scale up the values of certain parameters (e.g., perceived severity) but would not alter the qualitative feedback dynamics we study. The effects of waning immunity depend on its severity. If prior infection continues to reduce morbidity and mortality, we would not expect substantial deviations from our findings. In contrast, if immunity wanes rapidly and completely, it is conceivable that even strict lockdowns could remain in place for the entire duration of the simulation due to sustained transmission. While these extensions are important for future, more biologically detailed models, we chose to exclude them here to maintain focus on the core feedbacks between epidemic dynamics, health opinions, and interventions.

In recent years, epidemiological models that account for interaction between epidemics, health perceptions and behaviors have received growing attention, with SARS-CoV-2 pandemic bringing this synergetic relationship to the forefront [ [24–28] and many therein]. Numerous mathematical studies have shown that to accurately reconstruct observed epidemic dynamics, mathematical model should inlcude endogenous behavioral feedback which arises due to risk perception, peer communication, public media information dispersal and fatigue due to public health inteventions aiming to decrease onward pathogen transmission [29–31]. The relative simplicity in modeling infection transmission and opinion dynamics in our model was intentional, as our focus was on qualitative rather than quantitative analysis. Several studies using coupled epidemic-opinion dynamics investigated the impact of lockdown interventions by using models that incorporate lockdown fatigue. Some of these models modelled fatigue as exogeneous process [24, 30], while others captured it as endogeneous process driven by the system’s dynamics [31]. Most studies sought to isolate the effect of the fatigue on the observed dynamics, while others explored hypothetical scenarios where by means of public media interventions fatigue was reduced. In our work, we sought to demonstrate how the interplay of lockdown measures, infection dynamics, and aspects such as adherence and public fatigue can influence overall epidemic trends. Our findings highlight that the interactions between these elements can explain unexpected and, sometimes, undesirable results, underscoring the challenges of identifying the most effective interventions. Excluding the interplay between disease transmission, public health interventions, and health attitudes can significantly alter predicted outcomes. For example, Mellone et al. [32] employed a deterministic mathematical model to assess how the severity and length of lockdowns affected epidemic trends in Germany and Israel. Their model accounts for variations in social interactions during lockdowns but does not capture a dynamic response of public behavior adjusts to the intensity and extent of lockdown measures. Consequently, the forecasts by Mellone et al. differ from ours, suggesting that less stringent lockdowns with extended periods can also yield favorable results. In contrast, studies which included lockdown fatigue have demonstrated that in various settings, stringent and brief lockdowns might the optimal strategy to mitigate an outbreak (e.g., [24, 30].

This work demonstrates the importance of incorporating health-opinion-epidemic feedback dynamics into epidemic models, as the success of public health measures depends not only on their design, but also on their sustained support within the population. Our findings offer a mechanism to explain how interventions that appear sound in principle, may nevertheless underperform if they trigger opinion shifts or fatigue that reduce adherence over time.

## Conclusion

Changes in a population’s attitude and adherence towards lockdown measures may greatly influence their success in reducing case numbers. More stringent, but shorter lockdowns may be more effective in preserving a positive attitude of the population towards taking protective measures and simultaneously reducing the economic burden of the measures. Concerning the timeliness of measures, a balance has to be found between early onset of measures preventing exponential growth and late onset which may have more support in the population at the time of implementation.

## Data Availability

Code used for generating the results is openly available online at https://github.com/JorenB/infections-lockdowns-opinions

## Supporting information

**S1 p. Technical details of simulation model.**

**S2 p. Parameter calibration.**

**S3 p. Additional results.**

## Acknowledgments

This research was supported by a grant from the Netherlands Organization for Health Research and Development (ZonMw, grant number 91216062). H.N. and M.K. acknowledge funding by the German Federal Ministry for Education and Research for the infoXpand project (031L0300A). The authors would like to thank Emil Iftekhar, Daphne Van Wees, Ganna Rozhnova, Mui Thi Pham, and Sophie Diexer for their feedback during the base model development, and Michiel van Boven and Martin Bootsma for valuable comments on the setup of the model and the interpretation of the results.

