## Supplementary information for "Impact of adherence and stringency on the effectiveness of lockdown measures: a modelling study"

#### S1 Appendix.

**Technical details of simulation model.**

##### Overview

The model captures the dynamics of infection spread and health opinion competition in a fixed-size population. Infection transmission dynamics is described by Susceptible-Infected-Removed (SIR) framework whereupon each individual belongs to one of three classes. Susceptible individuals who come into contact with infectious individuals can become infected. If transmission of infection has taken place, individuals become infectious. After a period of time infectious individuals recover and become immune to further acquisition of infection. There are two mutually exclusive health opinions circulating in the population: health-positive, $\bigoplus$, and health-neutral, $\bigotimes$. Each individual holds one of two opinions. Individuals can switch opinion upon interacting with their peers who hold the opposite opinion. The more such individuals the faster this switch will occur. Switching of opinions can occur in a disease-free population.

Infection transmission dynamics and opinion switching are coupled via two mechanisms. Presence of the infection as captured by the global prevalence leads to increase in the switch rate to health-positive opinion. If in a response to the outbreak a lockdown is initiated, switch rate to health-neutral opinion increases, such that the increase is positively correlated to stringency and the duration of the lockdown.

Below we describe mathematical formulations and methods used to simulate the described dynamics.

##### Networks

In the context of the model definition, individuals are represented as nodes in a multiplex network. The first layer of the network (physical network) corresponds to

**Table A. Model parameters**

| Symbol | Description | Value(s), unit |
|---|---|---|
| $\beta, K$ | Watts-Strogatz algorithm [1] rewiring probability and mean degree, respectively | $\beta = 0.08$<br>$K = 14$ |
| $\gamma$ | Disease recovery rate | 1.0, week$^{-1}$ |
| $c_{\text{phys}}(j, j')$ | Physical contact rate between individuals $j, j'$ | 7.0, week$^{-1}$ |
| $c_{\text{inf}}(j, j')$ | Information contact rate between individuals $j, j'$ | 10.0, , week$^{-1}$ |
| $p$ | Opinion switch probability per information contact event | 0.04 |
| $k, \theta$ | Opinion switch propensity function shape parameters | $k = 1.8$<br>$\theta = 7.0$ |
| $\epsilon_\oplus, \epsilon_\otimes$ | Infection probability for opinion $\bigoplus$, $\bigotimes$ individuals per physical contact event with infected peer (of arbitrary opinion) | $\epsilon_\oplus = 2.0 \times 10^{-2}$<br>$\epsilon_\otimes = 3.5 \times 10^{-2}$ |
| $C_{\text{fat}}$ | Weight factor of lockdown fatigue contribution to opinion switch propensity | $5.0 \times 10^{-2}$ |
| $C_{\text{hs}}$ | Weight factor of prevalence related contribution to opinion switch propensity | 2.0 |
| $f_s$ | Threshold prevalence for lockdown initiation | $(0.5, 1.0, 2.0, 3.5, 5.0) \times 10^{-2}$ |
| $f_e$ | Threshold prevalence for lockdown lifting | $1.5 \times 10^{-3}$ |
| $q$ | Lockdown stringency | 0.00, 0.25, 0.5, 0.75, 1.00 |
| $\alpha$ | Lockdown adherence for opinion $\bigotimes$ individuals (high, medium, low) | 1.0, 0.5, 0.0 |

physical interactions which can result in infection acquisition. The second layer (social network) represent social interactions necessary for opinion competition dynamics.

In each network, connection of two nodes($j$ and $j'$) by an edge represents an interaction that persists in time with a certain frequency. In the physical network the frequency of interactions (physical contact rate) is denoted by $c_{\text{phys}}(j, j')$, in social network (information contact rate) it is denoted by $c_{\text{inf}}(j, j')$.

At the start of the simulation, both networks are identical. However, as an outbreak develops and lockdowns are initiated, the physical network changes its structure. Once a lockdown is relaxed, the physical network reverts to its pre-lockdown state. The networks are small-world networks created using the Watts-Strogatz algorithm [1], which starts with a ring lattice with average degree $K = 14$ and rewires network edges with probability $\beta = 0.08$ per edge. The resulting networks have clustering coefficient 0.54 and average path length 5.4.

We present a schematic representation of a small-world population network in Fig. S1. The label of a node indicates the disease state of an individual as either Susceptible (S), Infected (I), or Recovered (R). The color of the node indicates the opinion status, being either health-positive (blue) or health-neutral (red). Solid lines mean that the contact edge between two individuals is of both a *physical* and *informational* nature, whereas a dashed line indicates that the physical nature of the edge is switched off temporarily due to an active lockdown state. Disease transmission can only occur through solid lines.

#### Simulation algorithm

The dynamics of infection transmission and opinion switching processes are modeled using the Gillespie Algorithm [2,3] as follows.

The state of the population, denoted $X$, which encompasses both epidemiological state distribution and opinion distribution, can be modified by an event $l$. This event

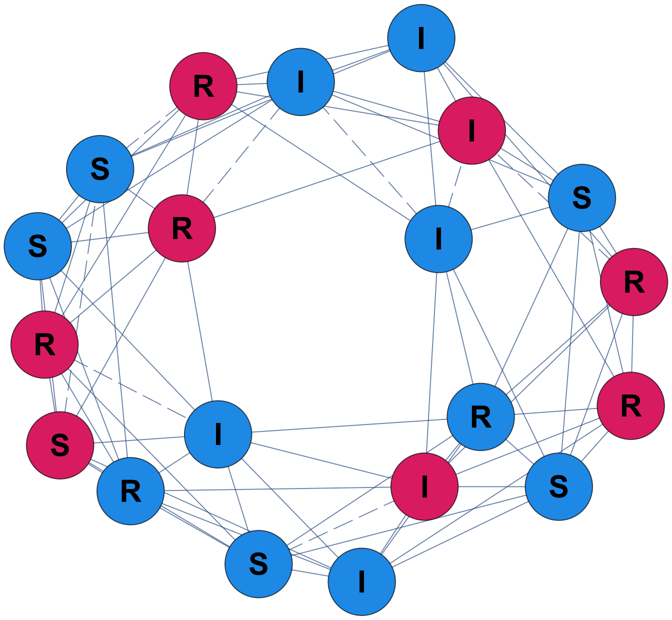

**Fig S1. A small-world network consisting of 20 nodes.** In the example shown, the mean degree is $K = 4$ and the rewiring probability for the Watts-Strogatz algorithm is $\beta = 0.1$. Letters indicate disease status (Susceptible, Infected, or Recovered), and colors represent opinion state (blue for health-positive, red for health-neutral). Solid lines allow for both informational and physical contact, whereas dashed lines allow for informational contact only, preventing disease transmission through such an edge.

might either cause a change in individual's epidemiological status or a switch of their opinion. Each event has an associated propensity function $\phi_l(X)$ defined as follows. At a time $t$, given the state of the system $X(t)$, an event $l$ has probability of occurring in the interval $[t+\tau, t+\tau+\Delta\tau)$ given by

$$p(\tau, l|X(t)) = \phi_l(X(t)) \exp(-\phi_0 (X(t)) \tau)\Delta\tau \tag{S1}$$

where

$$\phi_0(X(t)) = \sum_l \phi_l(X(t)) \tag{S2}$$

where $l$ iterates over the set of all possible events.

Thus, at each time point, given the current state of the system, we calculate the propensities for each possible event to take place. Then, using the joint probability distribution given by Eq. (S1) and Eq. (S2), an event that occurs and the time of its occurrence are determined. The system is updated accordingly and the step is completed. Note that given a model population size $N$, in each step we need to calculate propensities for $2N$ events.

#### Epidemiological state switch propensities

In the context of infection transmission dynamics, each individual can experience one of two possible events: infection $(S \to I)$ or recovery $(I \to R)$ with transition rates $\phi_{SI}$ and $\phi_{IR}$, respectively. Note that at any given time, an individual can undergo either one or the other event.

The propensity of individual $j$ holding opinion $\mathrm{Op}(j) \in \{\bigoplus, \bigotimes\}$ with the set of infected peers on the physical interaction network given by $\mathrm{IPN}(j)$ to experience event

$S \to I$ is defined by

$$\phi_{SI}(j) = \epsilon_{\mathrm{Op}(j)} \sum_{j' \in \mathrm{IPN}(j)} c_{\mathrm{phys}}(j, j'), \tag{S3}$$

where $\epsilon_{\mathrm{Op}(j)}$ is the probability for infection acquisition per contact with an infected peer. The infection acquisition probability $\epsilon_{\mathrm{Op}(j)}$ is lower for individuals holding health-positive opinion $\bigoplus$ than for individuals with health-neutral opinion $\bigotimes$, and these probabilities remain fixed throughout the simulation.

Contact rates $c_{\mathrm{phys}}$ are a subject to presence of the lockdown. During a lockdown, a proportion $q \in [0, 1]$ of contact rates will be set to zero. This implements the effect of lockdowns on the contact rates and subsequently on transmission of infection. When the lockdown ends, the contact rates revert to their original value.

In the model, health-neutral individuals can partially ignore lockdown rules, so the probability that contact rates between two such individuals reduce to zero is smaller than for contact rates between people where at least one of the two peers is of health-positive opinion. We define the parameter $\alpha \in [0, 1]$ to encode the resistance of health-neutral individuals to lockdown rules, such that the lower is the value of $\alpha$ the less likely individuals to comply with lockdown.

Therefore, to calculate contact rates (and thus, the propensities of infection transmission) in the conditions of lockdown, we iterate through all edges $jj'$ of the physical interaction network, and set the contact rate (weight) of the edge to zero with probability $p_{jj'}$, which we define as

$$p_{jj'} = \begin{cases} (1-\alpha)q, & \text{if } \mathrm{Op}(j) = \mathrm{Op}(j') = B \\ q, & \text{otherwise} \end{cases} \tag{S4}$$

The $I \to R$ transition only depends on the recovery rate $\gamma$, and is simply written

$$\phi_{IR}(j) = \gamma. \tag{S5}$$

##### Opinion state switch propensities

Opinion switches can take place from $\bigoplus$ to $\bigotimes$ and from $\bigotimes$ to $\bigoplus$. The model incorporates opinion switching in the disease-free regime based on information exchange with peers of opposite opinion, as described in [4]. For an individual $i$, denote the fraction of their peers with health opinion $l \in \{\bigoplus, \bigotimes\}$ by $n_l(j)$. Then the rates with which individuals switch their opinion are given by the following functions

$$\tilde{\phi}_{\bigoplus\bigotimes}(j) = c_{\mathrm{inf}} \frac{p_{\bigotimes} n_{\bigotimes}(j)^k}{1 + \theta_{\bigotimes} n_{\bigotimes}(j)^k}, \quad \text{from } \bigoplus \text{ to } \bigotimes \tag{S6}$$

$$\tilde{\phi}_{\bigotimes\bigoplus}(j) = c_{\mathrm{inf}} \frac{p_{\bigoplus} n_{\bigoplus}(j)^k}{1 + \theta_{\bigoplus} n_{\bigoplus}(j)^k}, \quad \text{from } \bigotimes \text{ to } \bigoplus \tag{S7}$$

where $p_l$ is the probability of switching opinion per contact and $\theta_l$ and $k$ are parameters that determine the shape of the switch rate function. By adjusting these shape parameters, we can continuously transform the switch rate propensity from a linear dependence on $n_l$ to a function that saturates at a given threshold, and finally to a sigmoid shape. We present examples of several possible choices of the shape parameters in Fig. S2. In the simulations, we have set $k$ and $\theta$ such that switch rate function have sigmoidal shape which results in a possibility of stable co-existence of two opinions in the population. Probabilities of switching opinion per contact, $p$, were set to the same value for both opinions, and the initial proportion of individuals holding either opinion was set to 0.5. These settings, in the absence of infection, lead to a quasi-steady

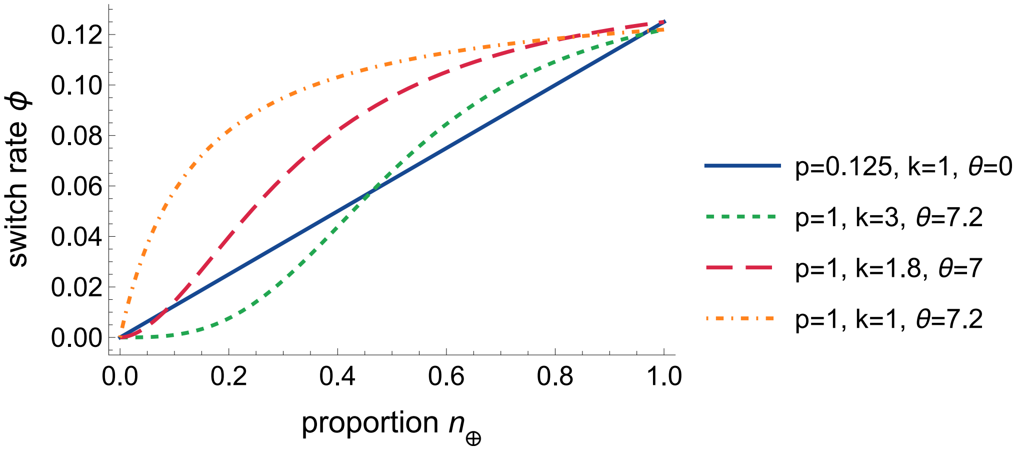


**Fig S2. Examples of opinion switch propensity functions.** By varying the shape parameters, we can model linear (blue, solid), saturating (orange, dash-dotted), and sigmoidal (red/green dashed) dependence of switch propensities on the independent variable. The green dashed sigmoid has a delayed growth onset as compared to the red dashed sigmoid. In our simulations, we have used function whose shape is similar to the red dashed curve.

distribution of opinions of around $(0.5, 0.5)$. Note that for simplicity we have implicitly assumed information contact rates of all edges to take the same value $c_{\text{inf}}$.

To model feedback relationship between infection spread, measures aiming to control it, and opinion competition, we modified the opinion switch propensity functions by including additional terms. To reflect the 'lockdown-fatigue' effect, which mimics people gradually getting weary of being in a lockdown regime and therefore, becoming more inclined to switch to a health-neutral opinion, we introduced a second term in the $\bigoplus \rightarrow \bigotimes$ opinion switch propensity function. We assume that the fatigue effect is stronger for more stringent and longer lockdowns. To capture this effect in the model we define a quantity $\xi_{\bigoplus\bigotimes}(t_l, q)$, where $t_l$ is the duration of the lockdown at time $t$ (i.e., the time since initiation of the lockdown measures until the current point $t$) and $q$ is the lockdown stringency. We assume that $\xi$ is an increasing function of both $t_l$ and $q$. In the simulations presented here, we defined $\xi$ as

$$\xi_{\bigoplus\bigotimes}(t_l, q) = C_{lf}\, t_l\, q. \tag{S8}$$

The parameter $C_{lf}$ determines the relative weight of the lockdown-fatigue function with respect to the opinion switch based on local opinion distributions, $\tilde{\phi}$.

Combining the two processes yields the propensity $\phi_{\bigoplus\bigotimes}$ for an individual $j$ with opinion $\bigoplus$ to change their opinion to $\bigotimes$ as follows:

$$\phi_{\bigoplus\bigotimes}(j) = \tilde{\phi}_{\bigoplus\bigotimes}(j) + \xi_{\bigoplus\bigotimes}(t_l, q). \tag{S9}$$

Similarly, to model the 'health-scare' dynamic, we introduce an additional term in the $\bigotimes\rightarrow \bigoplus$ opinion switch rate. 'Health-scare' refers to the phenomenon when people are influenced to adopt a health-positive opinion when the population-level disease prevalence increases. Therefore, during an outbreak the switch propensity $\bigotimes\rightarrow \bigoplus$ depends on the global disease prevalence $P$, with the functional dependence of the propensity for a health-scare-induced opinion given by

$$\xi_{\bigotimes\bigoplus}(P) = C_{hs}P. \tag{S10}$$

The complete expression for propensity $\phi_{\otimes\oplus}$ for an opinion switch from $\bigotimes$ to $\bigoplus$ is then

$$\phi_{\otimes\oplus}(j) = \tilde{\phi}_{\otimes\oplus}(j) + \xi_{\otimes\oplus}(P). \tag{S11}$$

### S2 Appendix.

**Parameter calibration.** The values of the infection probabilities $\epsilon_{\oplus}, \epsilon_{\otimes}$ and the opinion switch weight factors $C_{\mathrm{fat}}, C_{\mathrm{hs}}$ were fixed sequentially using a calibration procedure. For each parameter, we outlined heuristic conditions that we wanted our model to satisfy. We then performed 500 simulation runs for a range of candidate values of that parameter, and picked the value for which the appropriate heuristic condition was best met.

The first parameter to be calibrated was the probability of infection for health-positive individuals $\epsilon_{\oplus}$. The rationale for choosing the value of this parameter was that the infectious disease introduced to a population consisting solely of health-positive individuals should, in most cases, stay slightly below the epidemic threshold, meaning that the effective reproduction number is below 1. This means that the epidemic does not take off, but goes extinct after a short time. In the calibration runs for this parameter, no lockdowns were imposed, so that all individuals remain in the $\bigoplus$ state (cf. (S7)). We used the values $\epsilon_{\oplus} \in \{0.015, 0.02, 0.025, 0.03, 0.035\}$ as the candidate range. As seen in Fig. S3, the choice $\epsilon_{\oplus} = 0.02$ leads to a scenario where the median prevalence is projected to be small with the upper 95% quantile indicating that even in the case of the largest expected outbreak the prevalence will not reach high values. The median epidemic size is roughly the same for $\epsilon_{\oplus} = 0.025$, but the 95% quantiles now show larger outbreaks. Therefore, we fixed the infection probability for health-positive individuals to $\epsilon_{\oplus} = 0.02$.

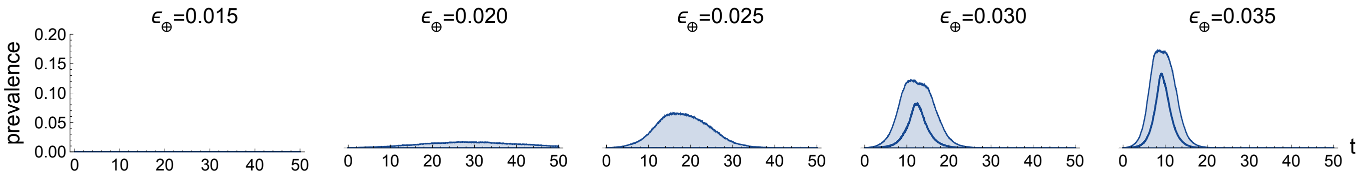


**Fig S3. Median and 95% CI of the prevalence over time ($t$, in weeks) for different values of the infection probability $\epsilon_{\oplus}$ for health-positive individuals.**

Next, we calibrated the probability of infection probability for health-neutral individuals $\epsilon_{\otimes}$, setting $\epsilon_{\oplus}$ to its fixed value 0.02 and the health-scare opinion switch weight factor $C_{\mathrm{hs}}$ to zero. We assumed that in a population with equal numbers of health-neutral and health-positive individuals larger outbreaks of the infectious disease should be possible. We tested the range $\epsilon_{\otimes} \in \{0.025, 0.03, 0.035\}$ and chose $\epsilon_{\otimes} = 0.035$ (see Fig. S4). Note that opinion switches do occur in these simulation runs, but since $C_{\mathrm{hs}} = 0$ and no lockdowns are imposed, the opinion switches are only induced by the local opinion distribution and the population-level opinion distribution remains approximately constant.

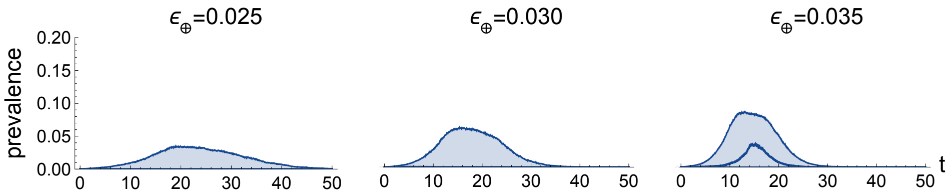


**Fig S4. Median and 95% CI of the prevalence over time ($t$, in weeks) for different values of the infection probability $\epsilon_{\otimes}$ for health-neutral individuals.**

The next parameter to be calibrated was the weight factor $C_{\mathrm{hs}}$ for the health-scare function $\xi_{\otimes\oplus}$ in the opinion switch propensity from health-neutral to health-positive (cf. equations S10 and S11). The rationale here was that the population should undergo a sizeable shift towards a health-positive opinion during an outbreak, but this should not lead to extinction of the health-neutral opinion. As shown in Fig. S5, the value $C_{\mathrm{hs}} = 2.0$ is a choice fulfilling this requirement. The rightmost graph shows that the health-neutral opinion goes extinct in the median scenario for the choice $C_{\mathrm{hs}} = 8.0$.

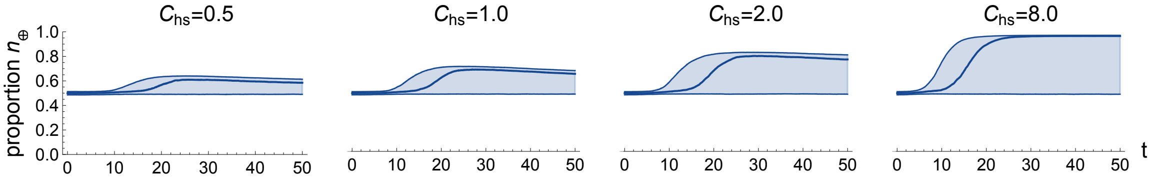


**Fig S5. Median and 95% CI of the proportion of health-positive individuals over time ($t$, in weeks) for different values of the health-scare weight factor $C_{\mathrm{hs}}$.**

Finally, we calibrated the weight factor $C_{\mathrm{fat}}$ for the lockdown fatigue function, which increases the propensity for health-positive individuals to switch to a health-neutral opinion (cf. equations S8 and S9). Using the previously described method to calibrate $C_{\mathrm{hs}}$, we chose a value of $C_{\mathrm{fat}}$ for which the population makes a strong shift towards a health-neutral opinion. The graphs shown in Fig. S6 support our choice of $C_{\mathrm{fat}} = 0.05$ for this parameter.

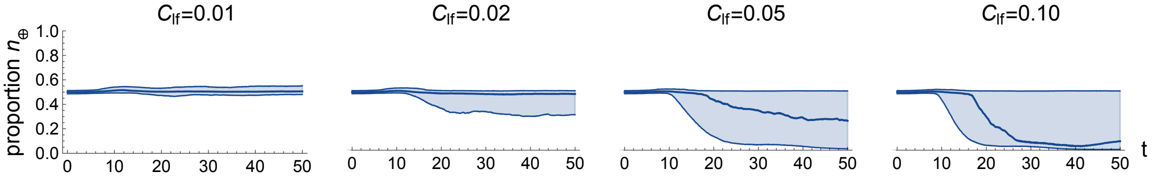


**Fig S6. Median and 95% CI of the proportion of health-positive individuals over time ($t$, in weeks) for different values of the lockdown fatigue weight factor $C_{\mathrm{fat}}$.**

### S3 Appendix

**Additional results.** In Fig. S7 we show the final proportion of health-neutral individuals (FPHN) for each individual simulation run, plotted against the total number of weeks during which there was an active lockdown in that run. This provides further insight into the mutual interaction between higher proportions of health-neutral individuals and higher disease prevalences. In this figure, the detrimential effect on the health opinion landscape of a non-effective lockdown is evident, regardless of whether it was due to low stringency or to low adherence. In this scenario, the lockdowns result in a significant increase in FPHN, not only compared to the baseline scenarios, but also compared to scenarios where lockdowns were more effective and subsequently shorter.

The results of additional sensitivity analyses for the stringency parameter $q$ and the adherence parameter $\alpha$ can be found on this project's GitHub page [5].

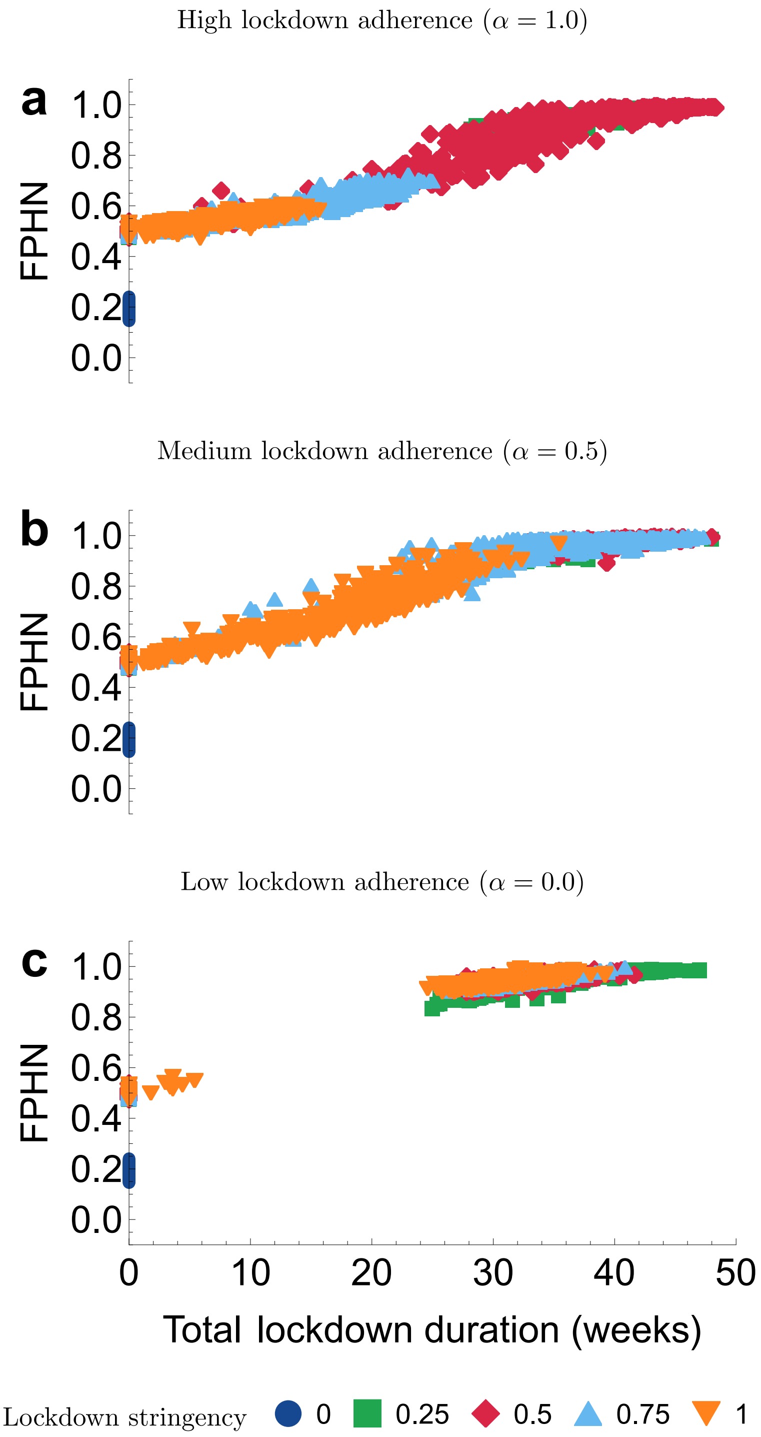


**Fig S7. Opinion distribution and lockdown duration.** The relative final proportion of health-neutral individuals (FPHN) versus the total lockdown duration, across various levels of lockdown adherence parameter $\alpha = 1.0, 0.5, 0.0$. The FPHN is reported relative to the mean FPHN in the scenario where no lockdown is implemented ($q = 0$). Each marker corresponds to a single simulation run. The threshold prevalence for lockdown initiation is fixed at $f_s = 0.005$.
